# Distance from tertiary care, pericardial effusion, and nutritional status predict all-cause mortality among Malawian children with rheumatic heart disease

**DOI:** 10.64898/2026.02.28.26346960

**Authors:** Jillian Olsen, Yamikani Chimalizeni, Jonathan Carapetis, Msandeni Chiume, Sarah Murphy Gunter, Mina Hosseinipour, Peter Kazembe, Subhrajit Lahiri, Treasure Mkaliainga, Kristy O. Murray, Daniel Penny, Tayamika Tambala, Anirudh Vinnakota, Amy Sanyahumbi

## Abstract

**Background:** This study of Malawian children with rheumatic heart disease (RHD) sought to detect demographic, clinical, and echocardiographic risk factors for mortality.

**Methods:** Pediatric patients with RHD were recruited from March to October, 2018 from clinic rosters and inpatient consults in Lilongwe and Blantyre, Malawi. An echocardiogram was performed upon study enrollment. Cox regression analyses were performed to assess for factors associated with mortality over nearly 2 years of follow-up.

**Results:** Of 118 patients, nearly two-thirds were female (64.4%) and median age was 12 (IQR 10-14). Just under half (47.0%) lived >40km from a tertiary care center. There was a high prevalence of severe mitral regurgitation (65.3%), and pericardial effusion was present in 18.6%. Nearly a quarter (23.7%) died during follow-up. In univariable Cox regression, living >40km from tertiary care, living in a remote area, moderate or severe malnutrition, taking a beta blocker, severe mitral stenosis, any severe valve disease, severe left atrial enlargement, and presence of a pericardial effusion were statistically significant risk factors for mortality (p<0.05). In the adjusted model, living >40km from tertiary care (HR 2.66, CI 1.06-6.07, p=0.037), malnutrition (mild HR 3.92, CI 1.03-14.91, p=0.045); moderate HR 7.41, CI 1.92-28.54, p=0.004; severe HR 4.91, CI 1.44-16.71, p=0.011), beta blocker use (HR 4.62, CI 1.63-13.10, p=0.004), and presence of a pericardial effusion (HR 6.96, CI 3.00-16.13, p<0.001) remained independent risk factors for mortality.

**Conclusions:** This study of Malawian children emphasizes the dire prognosis of RHD in under-resourced settings and provides potential area of focus for targeted intervention.

## 1. Introduction

While rheumatic heart disease (RHD) has been largely eliminated from developed nations, it represents the majority of acquired heart disease among young people globally^1^ and accounts for greater than 300,000 annual deaths worldwide. Reducing RHD is an integral part of World Health Organization (WHO) and World Health Federation (WHF) targets. In 2018, representatives from all six WHO regions unanimously adopted a Global Resolution on Rheumatic Heart Disease,^2^ which encourages high-quality research supporting “a broader understanding of the global disease burden.” Similarly, the World Heart Federation has called for a 25% reduction in RHD-related mortality among those younger than 25 years by 2025.^3^ As this deadline approaches, there remains a paucity of data about which patients are at highest risk of death, particularly among the pediatric population.

In the limited published studies available, mortality ranges widely from 2.9% of Fijian children with latent RHD detected on screening and followed for a median of 7.4 years^4^ to a staggering 31% of young people with clinical RHD followed for 8 years in Uganda.^5^ Moreover, there are incongruent findings on even the most basic risk factors for mortality such as sex and age, with some reporting female sex and younger age as protective^6^ and others finding the opposite.^7^ Increasing availability of echocardiography in traditionally under-resourced regions has permitted a greater understanding of the true burden of disease and may help detect those patients at risk of poor outcomes. This study seeks to identify demographic, clinical, and echocardiographic characteristics associated with mortality among Malawian children with RHD.

## 2. Methods

### 2.1. Study design and patient population

This was a prospective cohort study in which patients with a previous or new diagnosis of RHD were recruited from March to October, 2018 from outpatient and inpatient sources in Lilongwe and Blantyre, Malawi. Informed consent was obtained from a parent or guardian of each enrollee. Patients were followed until March 2020. Data were stored confidentially using REDCap database (Vanderbilt, Nashville, TN). The study protocol was approved by the Baylor College of Medicine Institutional Review Board (U.S.) and the National Health Sciences Research Committee (Malawi) and thus conforms to the ethical principles of the Declaration of Helsinki.

### 2.2. Data collection and definitions

Demographic and clinical data were collected from interviews and medical records, including age, age at time of RHD diagnosis, sex, distance between patient’s home and healthcare facilities, patient’s home setting (inner city, city suburb, large town, small town/village, remote area), number of people living in the home, family’s estimated monthly income, patient’s nutritional status, estimated severity of disease at time of diagnosis, whether the patient had a documented history of acute rheumatic fever, whether the patient underwent catheter-based or surgical cardiac intervention, and patient’s medications (penicillin, furosemide, aspirin, ACE inhibitor, digoxin, beta blocker, and warfarin).

Income was reported by each family in Malawi kwacha (Malawi currency) and was converted to U.S. dollars using the mid-market exchange rate on the patient’s day of study enrollment.^8^ Nutritional status was determined from age- and gender-based body mass index (BMI) z-scores according to the Centers for Disease Control 2000 growth charts.^9^ For the 2 patients who had weight but not height recorded, the weight z-score was used instead of BMI z-score. Anthropometric data at 1-year follow-up was used for one patient for whom enrollment height and weight were unavailable. Nutritional status was assigned as not malnourished for z-score ≥ −1, mildly malnourished for z-score < −1 and ≥ −2, moderately malnourished for z-score < −2 and ≥?-3, and severely malnourished for z-score < −3.^10^

Death occurring between study enrollment and the end of follow-up (March 2020) was the outcome variable. For patients whose date of death was imprecise (e.g. “shortly after diagnosis” or “April 2019”), date of death was assigned on the last day of the estimated month of death. The 8 patients for whom no approximate date of death was available were assigned March 5, 2020, the latest date on which follow-up data was gathered on any patient enrolled in the study, so as to not falsely exaggerate any factors associated with mortality.

An echocardiogram was performed on each patient at the time of enrollment in the study. Echocardiograms were interpreted by one of three readers in accordance with American Society of Echocardiography standards. Mitral stenosis was graded based on inflow gradient, pulmonary artery (PA) pressure, and valve area, where mild was assigned for gradient < 5mm Hg, PA pressure < 30 mmHg, and valve area > 1.5 cm^2^, moderate was assigned for gradient 5-10 mmHg, PA pressure 30-50 mmHg, and valve area 1-1.5 cm^2^, and severe was assigned for gradient > 10 mmHg, PA pressure > 50 mmHg, and valve area < 1 cm^2^.^11^

### 2.3. Statistical analysis

Each of the demographic and clinical characteristics was assessed as an independent variable in predicting survival, as measured by time from the enrollment echocardiogram to last follow-up or death. These variables included age, age at diagnosis, sex, distance from healthcare facilities, family size, home setting, income, nutritional status, severity of RHD at diagnosis, history of acute rheumatic fever, history of prior intervention, and medications. Echocardiographic features were likewise included as independent variables: severity of mitral regurgitation and stenosis, mitral valve inflow gradient, mitral valve area, severity of aortic regurgitation and stenosis, presence of severe disease of either the aortic or mitral valve (“severe valve disease”), involvement of both the aortic and mitral valves (“multiple valve disease”), presence of severe left atrial enlargement, left ventricular ejection fraction, and presence and size of pericardial effusion. After checking for proportional hazards, univariate Cox regression was performed, and those factors found to have a p-value < 0.1 for a risk of shorter survival were incorporated into a multivariable Cox regression. Backward elimination was used to arrive at the final model. For independent variables with a high degree of correlation or association (e.g. mitral stenosis severity and presence of severe valve disease), only the predictor with the smaller p-value was included in the model. Ordinal variables were treated as categorical. Statistical analysis was performed using SPSS IBM version 28.0 software.

## 3. Results

### 3.1. Patient population

A total of 124 patients were recruited initially. Six patients were ultimately lost to follow-up and were excluded from analysis, while the remaining 118 were followed for up to just under 2 years. Patient demographics and basic clinical data are reported in Table 1. Patients ranged in age from 5-18 years and were predominantly in early adolescence (interquartile range 10-14 years). Nearly two-thirds (64.4%) were female. About half of patients lived in remote areas and more than 40 km from the tertiary care center where they receive RHD care (either Kamuzu Central Hospital in Lilongwe or Queen Elizabeth Central Hospital in Blantyre). More than two-thirds of the families had more than 4 people, and family income was $1-$349 monthly (median $28, interquartile range $14-$69).

**Table 1.** Demographic and Clinical Characteristics by Vital Status.

|  | All (n=118) | Survived (n=90, 76.3%) | Died (n=28, 23.7%) | P-value* |
| --- | --- | --- | --- | --- |
| Age at Enrollment in Years, median (IQR) | 12 (10-14) | 13 (10-14) | 11 (10-13) | 0.103 |
| Age at Diagnosis in Years, median (IQR)<sup>†</sup> | 11 (8-13) | 10 (8-13) | 11 (9-13) | 0.193 |
| Female, n (%) | 76 (64.4) | 55 (61.1) | 21 (75.0) | 0.180 |
| Lives > 40 km from Tertiary Care Center<sup>‡</sup>, n (%)<sup>†</sup> | 55 (47.0) | 38 (42.2) | 17 (63.0) | 0.058 |
| Distance from Nearest Health Center, n (%)<sup>†</sup> |  |  |  | 0.415 |
| 0-10 km | 86 (73.5) | 68 (75.6) | 18 (66.7) |  |
| 11-20 km | 26 (22.2) | 19 (21.1) | 7 (25.9) |  |
| > 20 km | 5 (4.3) | 3 (3.3) | 2 (7.4) |  |
| Family Size > 4 People, n (%) | 83 (70.3) | 63 (70.0) | 20 (71.4) | 0.885 |
| Remote Residence (vs. Town, Suburb, or City), n (%) | 62 (52.5) | 42 (46.7) | 20 (71.4) | **0.022** |
| Monthly Income in US Dollars, median (IQR) | 28 (14-69) | 39 (14-69) | 21 (14-52) | 0.278 |
| Nutritional Status, n (%)<sup>‡</sup> |  |  |  | **0.049** |
| Not Malnourished | 50 (43.1) | 44 (48.9) | 6 (23.1) |  |
| Mildly Malnourished | 27 (23.3) | 21 (23.3) | 6 (23.1) |  |
| Moderately Malnourished | 19 (16.4) | 13 (14.4) | 6 (23.1) |  |
| Severely Malnourished | 20 (17.2) | 12 (13.3) | 8 (30.8) |  |
| RHD Severity at Diagnosis, n (%)<sup>¶</sup> |  |  |  | 0.259 |
| Mild | 5 (5.4) | 4 (5.6) | 1 (4.5) |  |
| Moderate | 8 (8.6) | 8 (11.3) | 0 (0.0) |  |
| Severe | 80 (86.0) | 59 (83.1) | 21 (95.5) |  |
| Prior Acute Rheumatic Fever, n (%) | 14 (11.9) | 12 (13.3) | 2 (7.1) | 0.514 |
| Has Undergone Catheterization or Surgery, n (%) | 11 (9.3) | 11 (9.3) | 0 (0.0) | 0.064 |
| Taking Penicillin, n (%) | 116 (98.3) | 89 (98.9) | 27 (96.4) | 0.420 |
| Taking Furosemide, n (%) | 102 (86.4) | 74 (82.2) | 28 (100.0) | **0.012** |
| Taking Aspirin, n (%)<sup>§</sup> | 41 (35.7) | 27 (30.7) | 14 (51.9) | **0.045** |
| Taking ACE Inhibitor, n (%)<sup>‡</sup> | 81 (69.8) | 61 (68.5) | 20 (74.1) | 0.583 |
| Taking Digoxin, n (%)<sup>‡</sup> | 2 (1.7) | 2 (2.3) | 0 (0.0) | 1.000 |
| Taking Beta Blocker, n (%)<sup>§</sup> | 13 (11.3) | 6 (6.9) | 7 (25.0) | **0.015** |
| Taking Warfarin, n (%)<sup>§</sup> | 6 (5.2) | 6 (6.9) | 0 (0.0) | 0.333 |

\*Reflects chi-square, Fisher's exact, or Mann Whitney U test as appropriate
N for each variable is 118 except where noted: <sup>†</sup>n=117; <sup>‡</sup>n=116; <sup>§</sup>n=115; <sup>¶</sup>n=93
<sup>¶</sup>Kamzu Central Hospital or Queen Elizabeth Central Hospital

Patients were enrolled between the time of RHD diagnosis to over 12 years following diagnosis. Only 11.9% of them had a history of acute rheumatic fever, and the vast majority (86.0% of the 93 for whom history about their diagnosis was available) had severe RHD at the time they were initially diagnosed. Just 5.1% had undergone a surgical or catheter-based intervention. Of the medications tracked, benzathine penicillin (98.3%), furosemide (86.4%), and ACE inhibitors (68.6% of 116) were the most commonly taken, while under 12% of patients were on a beta blocker, warfarin, or digoxin.

### 3.2. Echocardiographic data

Patients’ enrollment echocardiograms (Table 2) demonstrated a high prevalence of advanced disease, with 75.4% demonstrating severe regurgitation or stenosis of at least one valve (mitral or aortic), and half the patients (50.0%) having involvement of both the mitral and aortic valves.

**Table 2.** Echocardiographic Characteristics by Vital Status.

|  | All (n=118) | Survived (n=90, 76.3%) | Died (n=28, 23.7%) | P-value* |
| --- | --- | --- | --- | --- |
| Mitral Regurgitation Severity, n (%) |  |  |  | 0.082 |
| None/Trivial | 16 (13.6) | 15 (16.7) | 1 (3.6) |  |
| Mild | 12 (10.2) | 11 (12.2) | 1 (3.6) |  |
| Moderate | 13 (11.0) | 11 (12.2) | 2 (7.1) |  |
| Severe | 77 (65.3) | 53 (58.9) | 24 (85.7) |  |
| Mitral Stenosis Severity, n (%) |  |  |  | **0.002** |
| None/Trivial | 20 (16.9) | 19 (21.1) | 1 (3.6) |  |
| Mild | 21 (17.8) | 20 (22.2) | 1 (3.6) |  |
| Moderate | 43 (36.4) | 31 (34.4) | 12 (42.9) |  |
| Severe | 34 (28.8) | 20 (22.2) | 14 (50.0) |  |
| Mitral Stenosis Gradient (mmHg), median (IQR)<sup>§</sup> | 7.3 (4.7-10.8) | 6.7 (4.0-9.3) | 8.9 (6.9-12.4) |  |
| Mitral Valve Area (cm<sup>2</sup>), median (IQR) | 2.8 (2.0-3.9) | 2.8 (1.9-3.9) | 3.2 (2.3-4.0) |  |
| Aortic Regurgitation Severity, n (%)<sup>†</sup> |  |  |  | 0.473 |
| None/Trivial | 65 (55.6) | 51 (57.3) | 14 (50.0) |  |
| Mild | 6 (5.1) | 5 (5.6) | 1 (3.6) |  |
| Moderate | 28 (23.9) | 22 (24.7) | 6 (21.4) |  |
| Severe | 18 (15.4) | 11 (12.4) | 7 (25.0) |  |
| Aortic Stenosis Severity, n (%)<sup>†</sup> |  |  |  | 0.511 |
| None/Trivial | 109 (93.2) | 84 (94.4) | 25 (89.3) |  |
| Mild | 7 (6.0) | 4 (4.5) | 3 (10.7) |  |
| Moderate | 1 (0.9) | 1 (1.1) | 0 (0.0) |  |
| Severe Valve Disease, n (%) | 89 (75.4) | 63 (70.0) | 26 (92.9) | **0.014** |
| Multiple Valve Disease, n (%) | 59 (50.0) | 44 (48.9) | 15 (53.6) | 0.665 |
| Severe Left Atrial Enlargement, n (%)<sup>‡</sup> | 81 (70.4) | 55 (63.2) | 26 (92.9) | **0.003** |
| Left Ventricle Ejection Fraction <55%, n (%) | 9 (7.6) | 5 (5.6) | 4 (14.3) | 0.213 |
| Left Ventricle Ejection Fraction (%), median (IQR)<sup>†</sup> | 61.3 (56.9-65.0) | 62.0 (56.9-65.3) | 60.8 (57.1-64.1) |  |
| Pericardial Effusion Present, n (%) | 22 (18.6) | 8 (8.9) | 14 (50.0) | **<0.001** |
| Pericardial Effusion Size, n (%) |  |  |  | **<0.001** |
| None | 99 (81.4) | 82 (91.1) | 14 (50.0) |  |
| Trivial | 3 (2.5) | 0 (0.0) | 3 (2.5) |  |
| Small | 14 (11.9) | 6 (6.7) | 8 (28.6) |  |
| Moderate | 5 (4.2) | 2 (2.2) | 3 (10.7) |  |

\*Reflects chi-square, Fisher's exact, or Mann Whitney U test as appropriate
N=118 except where noted: <sup>†</sup>n=117; <sup>‡</sup>n=115; <sup>§</sup>n=110; <sup>||</sup>n=96

The mitral valve was most frequently affected, with 86.4% and 83.0% having at least mild mitral regurgitation and stenosis, respectively. Just under half of patients (44.4% of 117 with data on the aortic valve) had at least mild aortic regurgitation, while only 8 patients had at least mild aortic stenosis. Left atrial enlargement was also common, with 70.4% of 117 having severe left atrial enlargement. Just under one in five patients (18.6%) had a pericardial effusion. Despite the severity of valvar disease, left ventricular function was largely preserved, with 92.4% of patients having an ejection fraction > 55%.

### 3.3. Survival analysis

Almost a quarter (23.7%) of the children died during the two-year follow-up period. The mortality rate within 1 year following the diagnosis of RHD was also quite high at 11.0%. Based on the univariable Cox regression analysis (Table 3), living > 40 km from a tertiary care hospital, living in a remote area (as opposed to a town, suburb, or city), moderate or severe malnutrition, treatment with a beta blocker, severe mitral stenosis, any severe valve disease, severe left atrial enlargement, and the presence of a pericardial effusion were all statistically significant risk factors for mortality (p<0.05). Effusion size was also evaluated but was no longer associated with mortality when those patients without effusion were excluded, suggesting the presence not the size of effusion was associated with mortality. Age, sex, family size, and family income were not associated with death in univariable analysis.

**Table 3.**
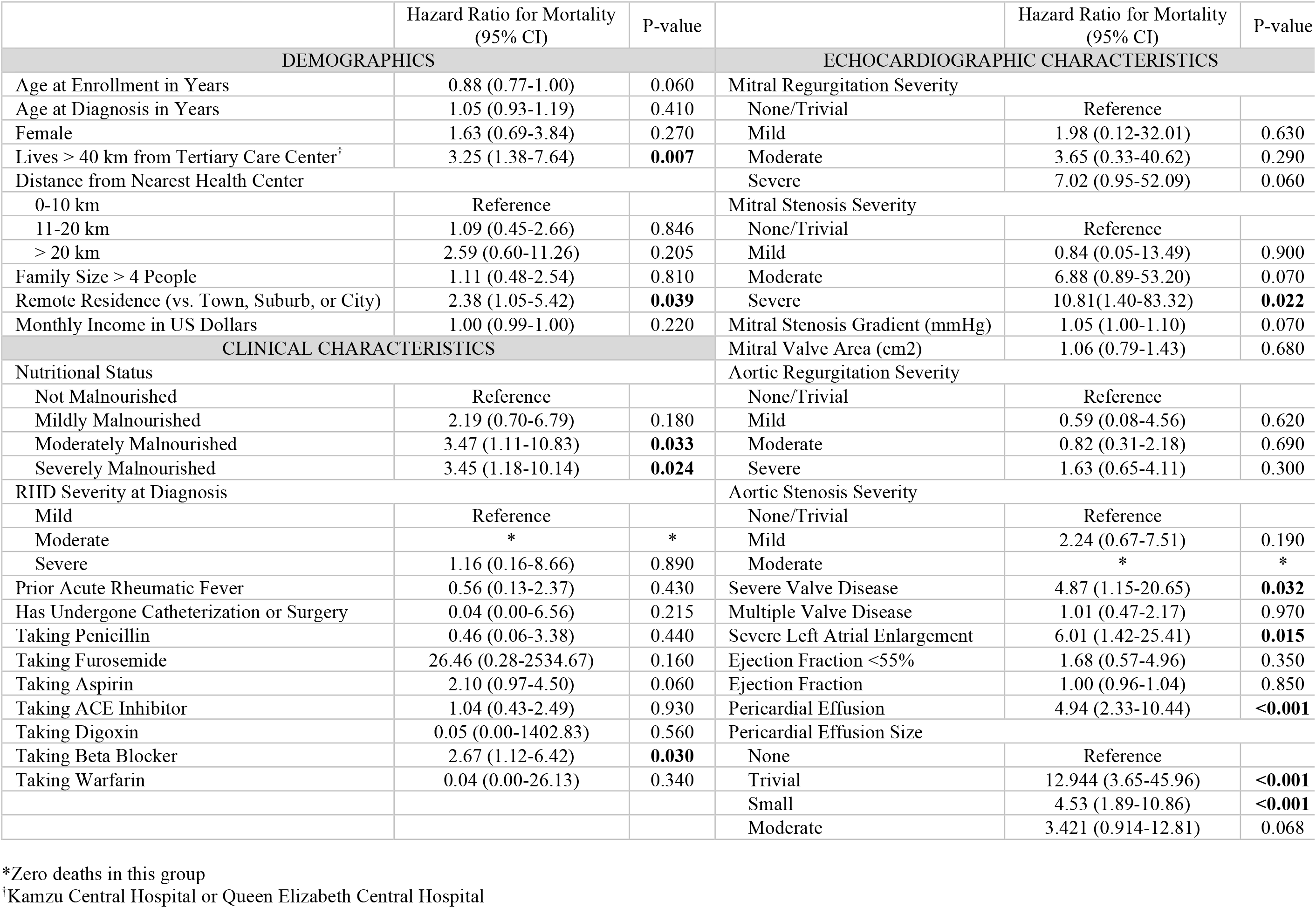
Univariable Cox Proportional Hazards Regression for Mortality.

In multivariable Cox regression models, living farther from tertiary care, greater malnutrition, being prescribed a beta blocker, and the presence of a pericardial effusion on initial echocardiogram remained independent risk factors for mortality in the follow-up period (> 40 km from tertiary care hospital HR 2.66, CI 1.06-6.07, p=0.037; mild malnutrition HR 3.92, CI 1.03-14.91, p=0.045); moderate malnutrition HR 7.41, CI 1.92-28.54, p=0.004; severe malnutrition HR 4.91, CI 1.44-16.71, p=0.011; beta blocker HR 4.62, CI 1.63-13.10, p=0.004, pericardial effusion HR 6.96, CI 3.00-16.13, p<0.001). Kaplan-Meier curves are presented in Figure 1 for visualization, and a map of patient homes is provided in Figure 2.

**Figure 1.**
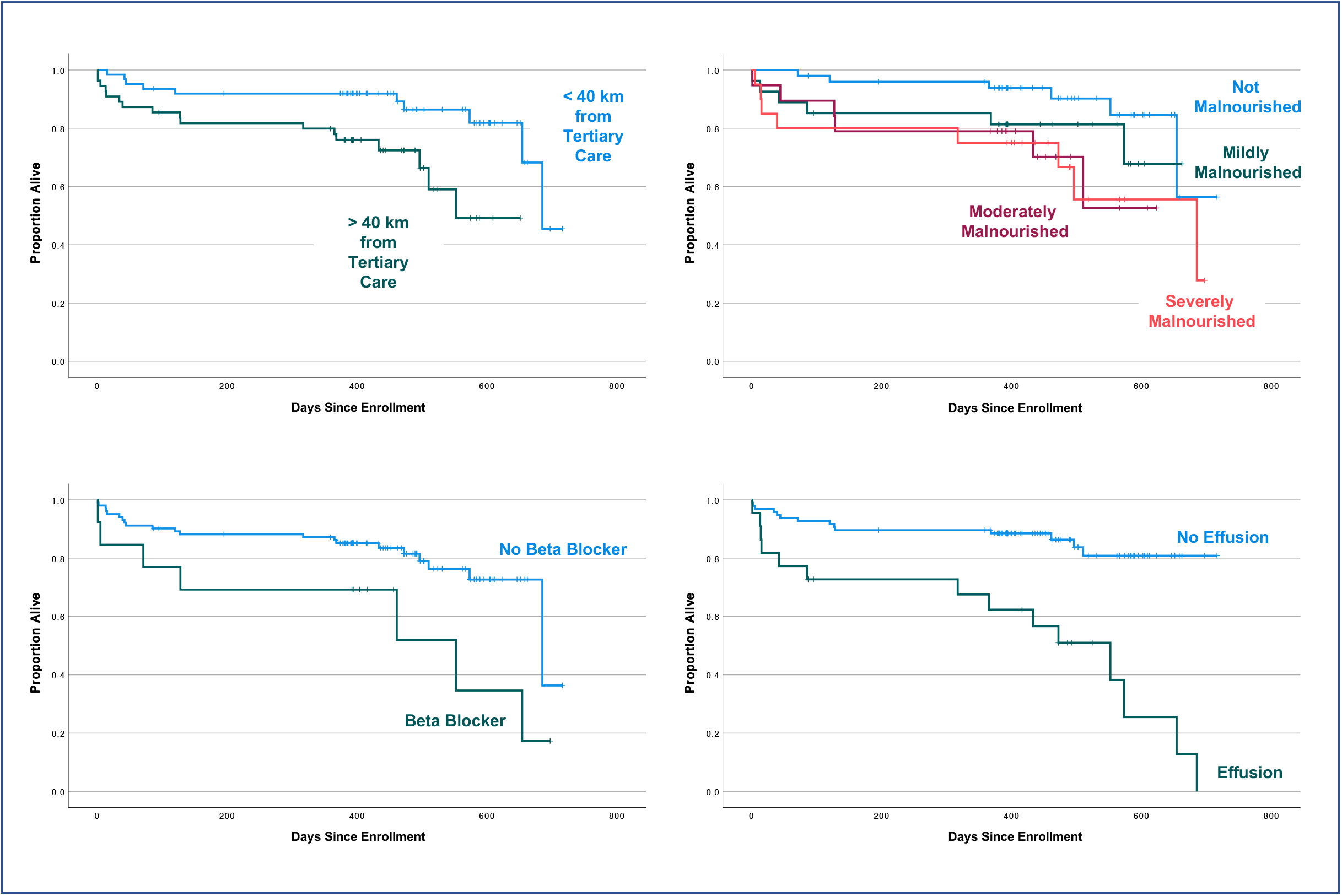
Kaplan-Meier Estimates of Survival by Distance from Tertiary Care, Nutritional Status, Beta Blockade, and Pericardial Effusion. *For 8 patients with unknown date of death, the last date of follow-up for any patient was assigned as the date of death

## 4. Discussion

This study of Malawian children with clinically diagnosed RHD emphasizes the dire prognosis of this disease, with more than 1 in 10 study participants dying within a year of diagnosis, and nearly 1 in 4 dying within 2 years of enrollment. Of course, these are the children who have been able to access medical care; there may be a significantly larger pool of unidentified cases and fatalities. The demographics of children with RHD in our study reflect those identified elsewhere, with a female predominance^12^ and peak age in early adolescence,^13^ making these results potentially generalizable to children with clinically-diagnosed RHD in other similarly under-resourced countries.

### 4.1. Echocardiographic characteristics

The markedly advanced illness found among our patients, with more than three-quarters having severe disease of at least one valve on initial echocardiogram, is consistent with other sub-Saharan African populations. Okello et al. showed that half of newly diagnosed RHD patients in Kampala, Uganda presented with severe mitral regurgitation on echocardiogram, and already had complications such as heart failure, pulmonary hypertension, atrial fibrillation, rheumatic fever recurrence, infective endocarditis, and stroke.^14^ Another hospital-based study in Cameroon likewise found that severe lesions were present in 80.9% of hospitalized patients of all ages with RHD,^15^ and 63.3% of pediatric patients similarly had severe valve involvement.^16^

However, while mitral regurgitation is commonly identified as the most prevalent lesion in RHD,^13,16,17^ this study demonstrated an unusually high proportion of stenotic mitral valves on initial echocardiogram. Stenosis is generally thought to develop later in the disease course, as reflected by greater frequency among adult patients with RHD.^13^ In other pediatric populations, well under 20% of patients have been found to have mitral stenosis, significantly less than the 80% identified here.^13,16,18^ The reason for this difference is not immediately apparent, though it may reflect overall later identification of disease, more frequent *Streptococcal* infections at an earlier age, or region-specific differences in disease patterns.

### 4.2. Mortality risk factors

RHD-related deaths in prior studies have been attributed to heart failure, stroke, atrial fibrillation, pulmonary hypertension, pregnancy related complications, and complications related to surgery.^15^ Factors previously associated with mortality include poor adherence to penicillin prophylaxis, heart failure, left ventricular dilation, severe valve disease, atrial fibrillation, lower education level, not having a surgical or catheter-based intervention, and residence in a lower or lower-middle income country.^5,6,14^ To our knowledge, this is the first study to demonstrate an association between mortality in pediatric RHD and malnutrition, pericardial effusion, beta blockade, or distance from tertiary care.

While chronic untreated heart disease could certainly result in malnutrition, explaining the noted association, it is likewise possible that the malnutrition contributes to disease progression. Both recurrent infection and an autoimmune response to infection play important roles in the pathogenesis of RHD, making immune system health a potentially critical factor in the severity of disease. Vitamin D deficiency has been correlated with both immune dysfunction^19^ and recurrent *Streptococcal* infections,^20^ and has been proposed as a causative mechanism in calcification of the mitral valve in RHD.^21,22^ It is feasible, then, that the association noted between malnutrition and death in this population could reflect this or other micronutrient deficiencies.

While effusions have been associated with mortality in many other disease processes,^23,24^ it is not yet clear why this is the case in RHD. The presence of a pericardial effusion on echocardiogram may reflect more severe valve disease, with resulting hypervolemia, risk of tamponade, or an inflammatory state. Regardless of the reason, this study suggests a need to prioritize urgent care for patients noted to have an effusion.

The association between beta blockers and mortality is likely reflective of the more frequent use of beta blockers in patients with worse mitral stenosis or arrhythmias rather than an inherent risk in their use. Indeed, all but 2 of the 13 patients taking beta blockers had severe mitral stenosis.

Although mitral stenosis did not remain statistically significantly associated with mortality on adjusted analyses, larger studies are needed to better define the relationship between valve disease, pericardial effusion, and clinical outcomes.

Intuitively, those with less access to specialized care fared worse. Previous studies have definitively linked greater distance from a health center with poor adherence to penicillin prophylaxis among adults and children with RHD,^25^ as well as with lower healthcare utilization for pediatric patients overall.^26^ The finding that in our study population, living farther from tertiary care carried a greater than 2-fold risk of mortality emphasizes the vulnerability of this population and the need for improved remote outreach and creative care delivery models.

Furthermore, none of the patients in our study who had undergone a catheter-based or surgical intervention died, and more than two-thirds of those who had a procedure lived < 40 km from tertiary care. Although not powered for statistical significance in our study, prior intervention was found to protective against mortality in a large pediatric Ugandan cohort.^5^ Improving access, especially to surgery for damaged valves, may be an important part of changing outcomes in RHD.

### 4.3. Study limitations

The recruitment of patients from both existing clinic patients and the newly diagnosed resulted in a study population that includes patients from all stages of disease. Thus, we are unable to draw conclusions about a patient’s echocardiogram or clinical status *at the time of diagnosis*. The interpretation of malnutrition as a risk factor for mortality is similarly limited by the collection of anthropometric data at the time of enrollment—we are unable to say whether preexisting malnutrition contributes to susceptibility to more aggressive disease, or whether malnutrition is a consequence of worse disease. Finally, the relatively small sample size and number of deaths limits the number of variables that can be included in the multivariable model with an expectation of generalizability. The study likewise may have been under-powered to identify additional potentially significant risk factors for mortality.

### 4.4. Conclusion

Severe valve disease is common among Malawian children with RHD, suggesting there is significant progress to be made in prevention, early identification of cases, and prevention of disease progression. Mortality rates are high, with 1 in 4 children in our cohort dying within 2 years of study enrollment. In resource-limited regions, it is crucial to be able to effectively identify those patients most likely to succumb to RHD in order to effectively distribute and target interventions. Special attention must be paid to those living far from tertiary care as this was an independent risk factor for mortality. Malnutrition and pericardial effusion on echocardiogram may also represent targets for triaging patients, treatment, or prevention, and should be explored further as the international efforts to curb the impact of RHD continue. If the World Health Assembly’s 2018 directive to change the course of RHD is not met with urgent action, the unnecessary deaths of children in countries like Malawi are likely to continue at devastating rates.

## Data Availability

All data produced in the present study are available upon reasonable request to the authors

